# Accelerated multi-organ proteomic aging is detectable decades before dementia onset

**DOI:** 10.64898/2026.01.16.26344299

**Authors:** Fangyu Liu, Wenyu Zhou, Aditya Surapaneni, Jingsha Chen, Cassandra Joynes, Pascal Schlosser, Zulema Rodriguez-Hernandez, Endre Sebestyén, Hamilton Se-Hwee Oh, Tony Wyss-Coray, Gabriela T. Gomez, Michael R. Duggan, Anna Prizment, Sanaz Sedaghat, Shuo Wang, Michael E. Griswold, Josef Coresh, Morgan E. Grams, Keenan A. Walker

## Abstract

Recent developments in proteomics have connected organ aging with dementia risk. The present longitudinal study extends this line of research by demonstrating that midlife organ age and *pace of organ aging* over multiple decades from midlife to late life are associated with future dementia risk and neurodegeneration, independent of the late-life organ age. We show further that advanced multi-organ aging, especially the combination of brain and heart/muscle aging, acts synergistically as a risk factor for dementia. Midlife proteome-wide analysis and Mendelian randomization identified a set of mostly non-brain-specific proteins driving or slowing the *pace of multi-decade brain aging*. Among these is tumor necrosis factor receptor superfamily member 1B (TNFRSF1B), causally implicated in the accelerated *pace of brain, immune, muscle, and pancreas aging*. Two other proteins associated with pace of brain aging, GM2 ganglioside activator (GM2A) and limbic system-associated membrane protein (LSAMP), showed putative causal roles in multiple neurologic diseases.

## Main

The geroscience hypothesis – that basic biological processes of aging may drive the risk of age-related diseases^1^ – has inspired research on biological aging clocks. Previously, human aging clocks have focused on the whole body.^2^ Recently, based on evidence suggesting that organs within an individual can age at different rates,^3^ measures of organ-specific age have been developed using clinical organ functions,^4^ DNA methylation,^5^ and plasma proteomics.^6,7^ The advent of the organ-specific age metrics enables the investigation of the link between organ-specific aging and age-related diseases, such as dementia, which have traditionally been considered a manifestation of biological disruptions to a single organ system.

Dementia, defined by significant cognitive impairment and functional decline, is a disease of the brain. However, health of the non-central nervous system (CNS) organs may contribute to the risk of dementia. For example, diabetes, cardiovascular disease, chronic obstructive pulmonary disease (COPD), and autoimmune diseases have each been associated with increased risk of dementia, implicating the pancreas, cardiovascular system, lungs, and immune system, respectively.^8–11^ Many studies of organ-specific aging have found that the plasma proteomics-based organ age of the brain and other organs, such as kidney, liver, and immune system, was associated with increased risk of all-cause dementia, Alzheimer’s disease (AD), and/or vascular dementia.^6,7,12–14^

Despite these recent developments, several important knowledge gaps remain, which limit our understanding of the organ-specific contributions to dementia risk. First, dementia may result from abnormalities of multiple organs, including but not limited to the brain. However, few studies have examined the potential synergistic effects of advanced age in multiple organs on dementia risk. Second, almost all studies to date have focused on a single measure of organ-specific ages. It remains unknown how longitudinal changes in organ age across life stages relate to dementia risk. Lastly, the biological processes and molecular drivers associated with rapid and resilient organ aging, and their potential direct and indirect effects on brain health, have not been fully investigated.

To address these gaps, we leveraged the plasma proteomic data in the Atherosclerosis Risk in Communities (ARIC) study, a multi-racial, population-based cohort (**Figure 1A)**.^15^ We found that older brain age in midlife and late life was associated with an elevated risk of subsequent dementia; although, older age in other organs (e.g., heart, immune system, liver, pancreas) had comparable or even stronger associations with dementia risk at both life stages. Additionally, we demonstrated that a faster *pace of organ aging* – defined as the rate of change in organ age between midlife and late life – in brain, intestine, muscle, and pancreas was associated with higher risk of dementia, independent of the late-life organ age, and we discovered many organ non-autonomous circulating proteins (i.e., proteins that are non-organ-specific or specific to other organs) associated with the *pace of aging* in brain and other dementia-associated organ systems, some of which were mechanistically implicated in neurologic disease.

**Figure 1.**
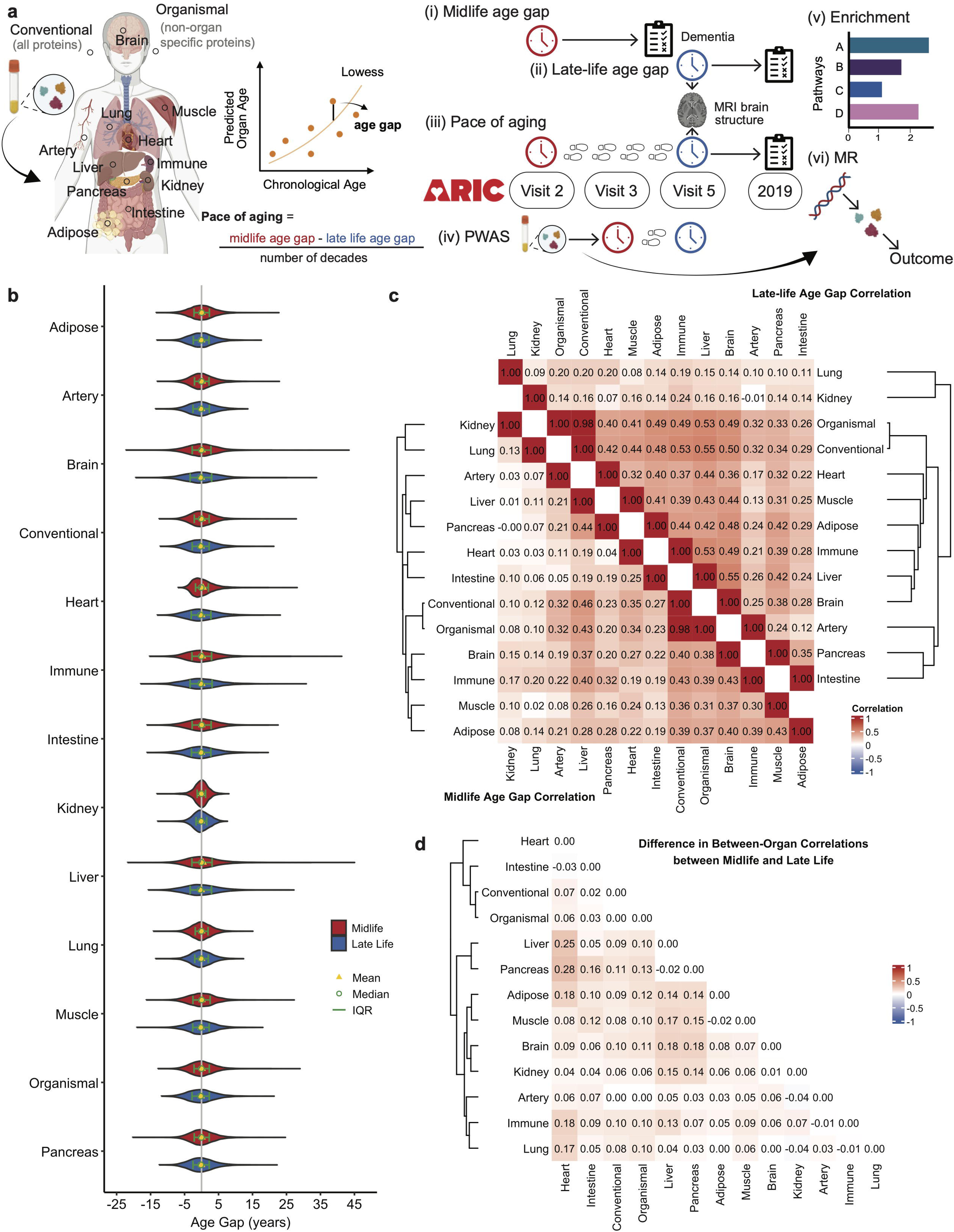
Study design and organ age gaps in midlife and late life. **a.** The organ age gaps were estimated using plasma proteomics and chronological age. The *pace of organ aging* was derived as the rate of change between midlife and late life (left). Primary statistical analyses related organ age gap (i and ii) and *pace of organ aging* (iii) to incident dementia using Cox proportional hazard models and MRI-based brain structure measures both using linear or logistic regression models (middle). Circulating proteins that may drive the *pace of organ age* was explored using linear regression models (iv, middle). The significant proteins were further annotated using enrichment analyses (v) and their causal link to *pace of organ aging* and neurologic disease examined using Mendelian randomization (vi, right). **b.** The violin plots show the distribution of organ age gaps in midlife and late life, respectively. **c.** The heatmaps display the correlation of age gaps between organs in midlife (lower) and late life (upper). Darker color indicates higher correlation. **d.** The heatmap depicts the change in between-organ correlations between midlife and late life. Darker color indicates greater change.

## Results

### Organ age gaps in midlife and in late life

Using the proteomics data from the ARIC study measured by the SomaScan assay^®^ (version 4.0), we estimated 13 biological ages using the algorithm developed by Oh and colleagues (Methods).^6^ Briefly, organ-specific proteins for 11 organ systems (adipose, artery, brain, heart, immune system, intestine, kidney, liver, lung, muscle, and pancreas) were identified as having gene expression in one organ at least 4 times higher than any other organ. Next, organ-specific proteins, non-organ-specific proteins (for organismal age), and all measured proteins (for conventional age) were trained to predict chronological age, respectively. We applied the weights from these prediction models in ARIC and estimated the organ-specific, organismal, and conventional ages at two visits: Visit 2, henceforth referred to as “midlife” (age 57.1±5.7 years, N = 11,595), and Visit 5, henceforth referred to as “late-life” (age 75.2±5.0 years, N = 4,287, **Figure 1A, Supplemental Table 1**). At each visit, the predicted biological ages were residualized using a locally weighted scatterplot smoothing (lowess) regression model (bandwidth of 2/3) between predicted biological ages and chronological age. These residuals are henceforth referred to as organ, conventional, or organismal “age gaps”. The age gaps had means close to 0 and standard deviations (SD) ranging from 2.1 to 5.0 years for midlife and 2.4 to 5.2 for late life (**Figure 1B**, **Supplemental Table 2**).

The correlations between the organ-specific age gaps within the same visit were weak to moderate, but the correlation tended to be greater during late life (mean r = 0.27) compared to midlife (mean r = 0.19, **Figure 1C, Supplemental Table 3**). This finding is consistent with previous literature and may suggest that each organ age gap captures a different dimension of biological age. Notably, there is a convergence of organ-specific ages over time; heart, liver and pancreas age show the greatest increases in their correlations with the other organ ages (**Figure 1D**).

We defined abnormal age as an age gap >1.5 SD and youthful age as an age gap <-1.5 SD. Participants with abnormal ages in dual organ systems comprised <2% of the midlife and late-life samples (**Supplemental Figures 1A and 1B**). Those with youthful ages in dual organs, or youthful age in one organ and abnormal age in another organ, were present in both the midlife and late-life samples, but the prevalence was extremely low (mostly <1%, **Supplemental Figures 1A and 1B**).

### Midlife and late-life organ-specific age gaps are associated with dementia

We first examined the associations of midlife and late-life organ-specific age gaps with the risk of incident dementia using Cox proportional hazard models adjusting for demographic (chronological age, sex, a composite variable of race and study center, and education) and physiologic factors (BMI, kidney function, current smoking status, history of diabetes and hypertension, and *APOE*ε4 status) known to jointly affect protein levels and dementia risk. In midlife (N=11,595), per-year higher (i.e., older) conventional and organismal age gaps had the strongest associations with incident dementia (949 cases) over a median 20.2-year follow-up period from Visit 2 to Visit 5 (**Figure 2A, Supplemental Table 4**). Of the organ-specific age gaps, per-year higher heart (HR=1.04; 95% CI: 1.02, 1.06) and brain (HR=1.04; 95% CI: 1.02, 1.05) age gaps were most strongly associated with incident dementia; artery, liver, intestine, and immune age gaps also showed significant (FDR p<0.05), albeit weaker, associations with incident dementia.

**Figure 2.**
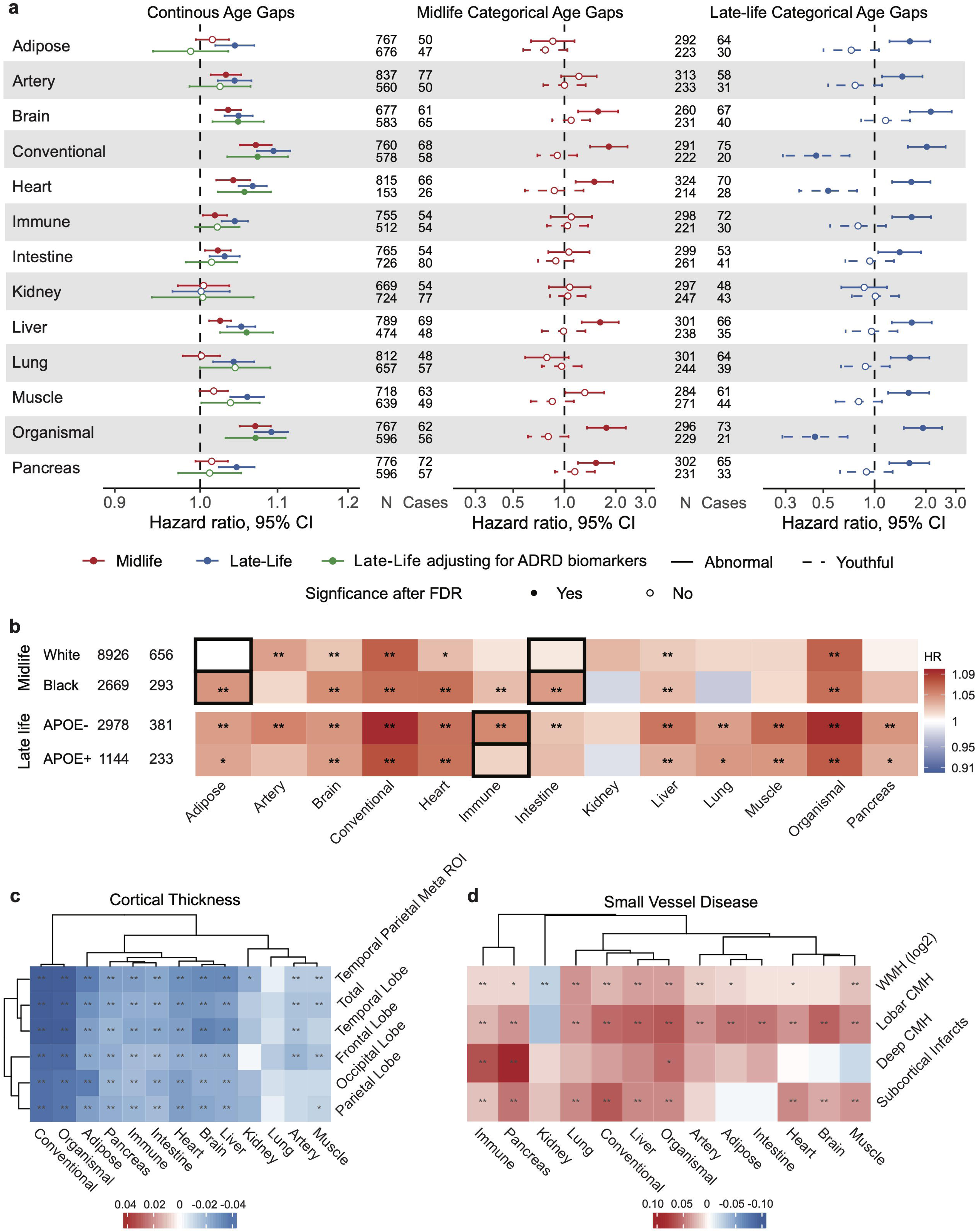
Organ age gaps were individually associated with incident dementia and MRI-defined cortical thickness and cerebral small vessel disease. All models were adjusted for chronological age, sex, a composite variable of race and study center, education, body mass index (BMI), kidney function (eGFR), current smoking status, history of diabetes and hypertension, and *APOE*ε4 status. **a.** The left forest plot shows the hazard ratios (HRs) for the associations with incident dementia, in midlife (red), late life (blue) and late life with adjustment of Alzheimer’s disease and related dementias (ADRD) biomarkers (green), when organ age gaps were modeled as continuous variables. The middle plot shows the HRs in midlife comparing individuals with abnormal (>1.5 SD) and youthful (<-1.5SD) organ age gaps to normal organ age gaps. The right plot shows the HRs in late life midlife comparing individuals with abnormal (>1.5 SD) and youthful (<-1.5SD) organ age gaps to normal organ age gaps. **b.** The HRs between organ age gaps and incident dementia comparing Black and White participants in midlife (upper) and comparing *APOE*ε4 carriers to non-carriers (lower). Black boxes indicate organ age gaps that showed significantly different associations with incident dementia between groups. **c.** The associations between organ age gaps and MRI-derived cortical thickness in 5 brain regions. Darker blue color indicates stronger association between more advanced organ age and lower critical thickness. **d.** The associations between organ age gaps and 4 MRI-derived small vessel disease measures. Darker red color indicates stronger associations between more advanced organ age and higher white matter hyperintensity (WMH, log2-transformed) or presence of cerebral microhemorrhage (CMH)/infarcts. ** indicates statistical significance at FDR level. * indicates statistical significance at nominal level.

In late life (N=4,287; 641 cases), higher age gaps in nearly all organs were significantly associated with incident dementia over a median follow-up of 6.5 years from Visit 5 to December 31, 2019 (FDR p<0.05). Conventional, organismal, and heart age gaps (HR=1.07; 95% CI: 1.05, 1.09) remained the strongest associations. Brain age gap (HR=1.05; 95% CI: 1.03, 1.07), however, ranked lower than muscle (HR=1.06; 95% CI: 1.04, 1.08) and liver (HR=1.05; 95% CI: 1.03, 1.07) in the magnitude of associations (**Figure 2A, Supplemental Table 5**). To assess reverse causation, i.e., preclinical dementia pathology affecting organ-specific age gaps, we removed 108 cases and 71 non-cases with follow-up time less than 3 years after Visit 5. We found slightly attenuated associations but unchanged significant results (**Supplemental Figure 2A**)

To further explore whether these biological age gaps capture dementia risk beyond that of widely used ADRD plasma biomarkers,^16^ we repeated the analyses adjusting for available measures of amyloid-*β* (A*β*42/A*β*40 ratio), phosphorylated tau 181 (p-tau 181), reactive astrogliosis (glial fibrillary acidic protein [GFAP]), and neuronal injury (neurofilament light chain [NFL]) at late life (Visit 5, N=1299, case=234). We observed minimal attenuation or even strengthening was observed for brain (HR=1.05; 95% CI: 1.02, 1.08), heart (HR=1.06; 95% CI: 1.02, 1.09), and liver (HR=1.06, 95% CI: 1.02, 1.09, **Figure 2A, Supplemental Table 6**).

Compared to normal age gaps (within 1.5 SD around the mean), *abnormal organ-specific age gaps* (>1.5 SD) during midlife in the brain (HR=1.58; 95% CI: 1.20, 2.07), heart (HR=1.50; 95% CI: 1.16, 1.93), liver (HR=1.62; 95% CI: 1.26, 2.08), and pancreas (HR=1.53; 95% CI: 1.20, 1.96) were associated with elevated risk of subsequent dementia, as was abnormal conventional and organismal age gaps. *Youthful organ-specific age gaps* (<1.5 SD) were not associated with lower dementia risk during midlife (**Figure 2A, Supplemental Table 7**). In late life, abnormal age gaps in all organs except kidney were associated with higher risk of dementia (HR ranging from 2.14 [brain, 95% CI: 1.61, 2.84] to 1.40 [intestine, 95% CI: 1.05, 1.88]), whereas youthful heart (HR=0.53; 95% CI: 0.36, 0.79), conventional, and organismal age gaps were associated with lower risk of dementia (**Figure 2A, Supplemental Table 8**).

### Race and genetics modify the link between organ-specific age gaps and dementia risk

As the algorithm for organ age gaps has so far been applied mostly to populations of European ancestry, we investigated whether the associations between the organ age gaps and dementia risk differed by race (White N=8,926; Black N=2,669). In late life, Black and White participants showed similar associations between organ-specific age gaps and dementia risk (p-interactions>0.05, **Supplemental Figure 3, Supplemental Table 10**). In midlife, however, adipose and intestine age gaps were more strongly associated with dementia risk among the Black, compared to White, participants (p-interactions<0.05, **Figure 2B, Supplemental Table 9**), suggesting that the early organ-specific dementia risk may differ by factors associated with race. Although racial differences in fat accumulation have been reported,^17–20^ the race-specific effect of adipose and intestinal aging on brain health outcomes remains largely underexplored.

We also examined whether *APOE*ε4 genotype (a major AD risk variant) and history of cardiovascular disease (diabetes, coronary heart disease, stroke, and heart failure) modified the associations between organ-specific age gaps and dementia risk. We observed greater associations between the late-life immune age gap and dementia risk among ε4 non-carriers compared to ε4 carriers (p-interactions<0.05, **Figure 2B, Supplemental Tables 11-12**), suggesting that advanced immune age may be more detrimental for non-AD dementia. History of cardiovascular disease did not modify the associations between organ-specific age gaps and dementia risk (p-interactions>0.05, **Supplemental Figure 3, Supplemental Tables 13-14**).

### Organ-specific age gaps are associated with structural brain metrics

Next, in a subset of 1,365 older adults without dementia (age 76.0±5.2 years) who had available MRI data, we examined how organ-specific age gaps relate to MRI-defined cortical thickness of the total brain, 4 lobes (frontal, parietal, temporal, and occipital lobes), and an AD-linked temporal parietal meta ROI.^21^ Mirroring the incident dementia results, per-year higher conventional and organismal age gaps had the strongest associations with lower cortical thickness in all regions after adjusting for demographic and physiological variables. Greater adipose, brain, heart, immune, intestine, liver, and pancreas age gaps also showed strong and widespread associations with reduced cortical thickness (**Figure 2C, Supplemental Table 15**). Similarly, immune, pancreas, and organismal age gaps showed the most consistent associations with the MRI measures of cerebral small vessel disease: greater white matter hyperintensity volume (N = 1,361), and the presence of lobar cerebral microhemorrhages (case/N = 251/1365), deep cerebral microhemorrhages (case/N = 51/1365), and subcortical infarcts (case/N = 254/1365, **Figure 2D, Supplemental Tables 16 and 17**). Surprisingly, higher brain and heart age gaps were strongly associated with higher odds of lobar cerebral microhemorrhages and cortical infarcts, but not with white matter hyperintensities or subcortical microhemorrhages, suggesting that these organ age gaps may be capturing processes linked to cortical and cardioembolic vascular injury, rather than the diffuse small vessel arteriopathy typically underlying WMH or deep microbleeds.

### Multiple organ age gaps demonstrated synergistic effects on dementia risk

Next, we examined whether the number of organs (excluding conventional and organismal) with abnormal age gaps (0, 1, 2, or 3+) during midlife and late life was associated with incident dementia. As depicted in **Figure 3A**, the risk of dementia did not increase linearly as the number of organs with abnormal age gaps increased. For example, having 2 organs with abnormal age gaps more than doubled the risk of dementia compared to having only 1 organ with an abnormal age gap in late life, though the deviation from linearity was not statistically significant (p>0.05).

**Figure 3.**
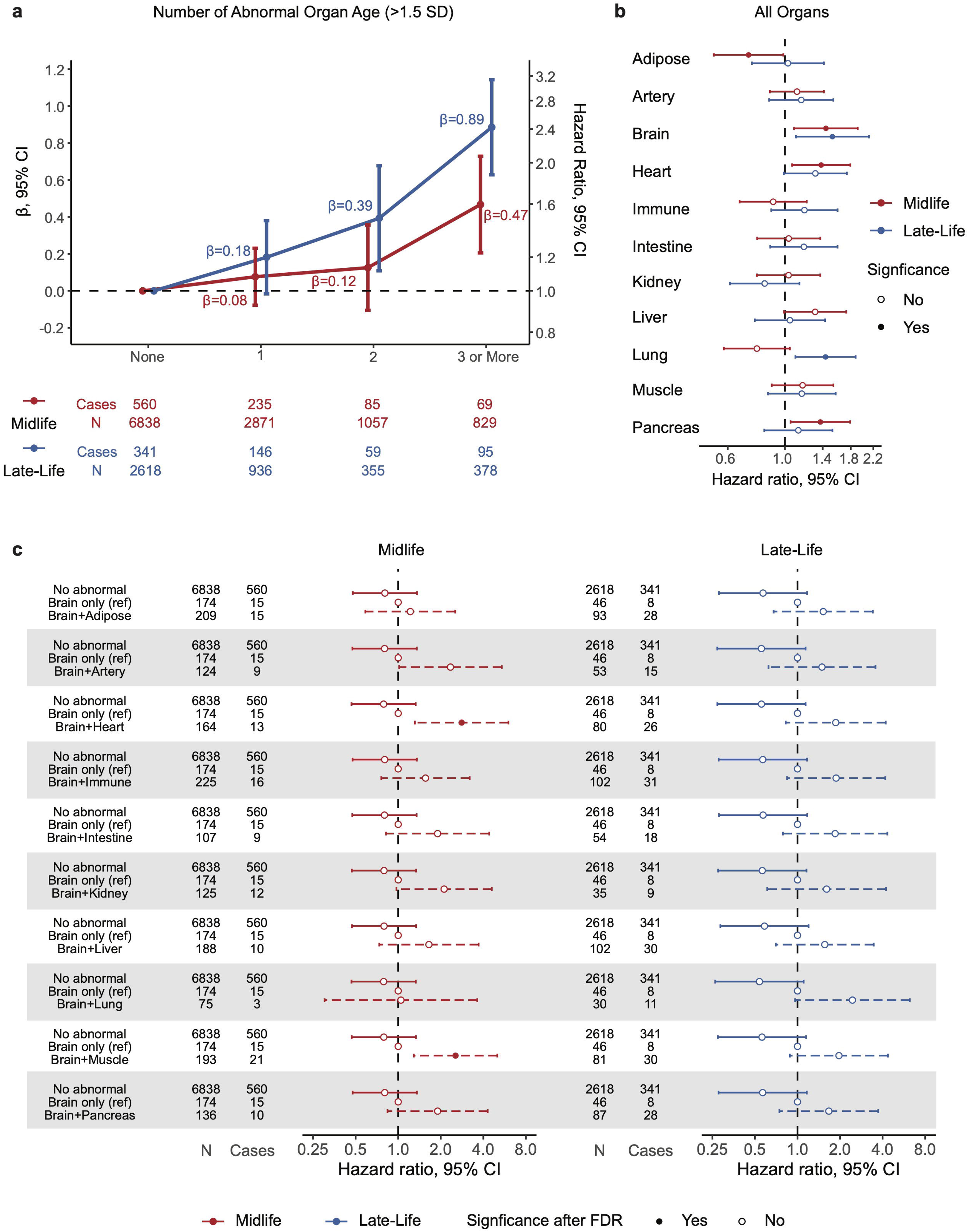
Advanced organ age gaps in multiple organs synergistically increased the risk of dementia. All models adjusted for chronological age, sex, a composite variable of race and study center, education, body mass index (BMI), kidney function (eGFR), current smoking status, history of diabetes and hypertension, and *APOE*ε4 status. **a.** The associations between the number of organ systems with abnormal age gaps (>1.5 SD) and incident dementia increased non-linearly in midlife (red) and late life (blue). **b.** The associations between age gap in each organ system and dementia risk in midlife (red) and late life (blue) after adjusting for the age gaps in all other organs and organismal age gap. **c.** The left forest plot shows the hazard ratios (HRs) of incident dementia comparing midlife individuals with normal age gaps (-1.5 SD to 1.5 SD) in all organ system and those with abnormal age gap (>1.5 SD) in brain and one other organ to those with abnormal age gap in brain only. The right plot shows the HRs of incident dementia comparing late-life individuals with normal age gaps (-1.5 SD to 1.5 SD) in all organ system and those with abnormal age gap (>1.5 SD) in brain and one other organ to those with abnormal age gap in brain only.

We then included 11 indicators of *abnormal age gap* in each organ and an indicator of *abnormal organismal age gap* in the same Cox model to determine which organ system was independently associated with dementia risk. In both midlife and late life, abnormal brain age gaps were associated with higher dementia risk after controlling for other age gaps (midlife HR=1.44; 95% CI: 1.08, 1.91; late-life HR=1.53; 95% CI: 1.10, 2.12, **Figure 3B, Supplemental Tables 18 and 19**), as were heart and pancreas age gaps in midlife and lung age gap in late life.

Because the brain organ age gap was independently associated with dementia risk at both life stages, we then examined how abnormal brain age, in combination with abnormal age gaps in other organs, may confer additional risk of dementia. For each organ system other than the brain, we categorized participants into groups defined by (i) no abnormal age gaps in any organ systems, (ii) abnormal age gap in brain only, and (iii) abnormal age gaps in brain and the organ system of interest. In midlife, compared to participants who had abnormal age gaps in the brain only, participants who had abnormal age gaps in brain and heart had significantly higher risk of dementia (FDR p<0.05, **Figure 3C, Supplemental Table 20**). A similar finding was observed for participants who had abnormal age gaps in brain and muscle. No combinations of the brain and another organ showed significantly higher risk of dementia in late life compared to abnormal brain age gaps alone (**Figure 3C, Supplemental Table 21**), likely due to limited power.

### Mid- to late-life pace of organ aging is associated with subsequent dementia risk and structural brain metrics

A total of 3,725 participants had predicted biological ages at both midlife and late life (average time interval of 20.7 years). The within-organ age gap correlations were moderate (0.4-0.6, **Figure 4A**), and over 50% of participants who had *abnormal age* in an organ system in midlife transitioned out of this category in late life (**Supplemental Figure 4**). While measurement error may explain some changes between visits, it may also suggest that proteomic measures of accelerated organ age are not static and may be amenable to intervention. We then calculated the difference between the late-life and midlife organ-specific age gaps and standardized the difference by the time interval (per decade), which we term the *pace of aging*. A positive value indicates faster organ-specific biological aging, compared to the population average rate of biological aging, over the same time interval. Hence, all paces of aging had a mean and median close to 0 (**Figure 4B, Supplemental Table 22**).

**Figure 4.**
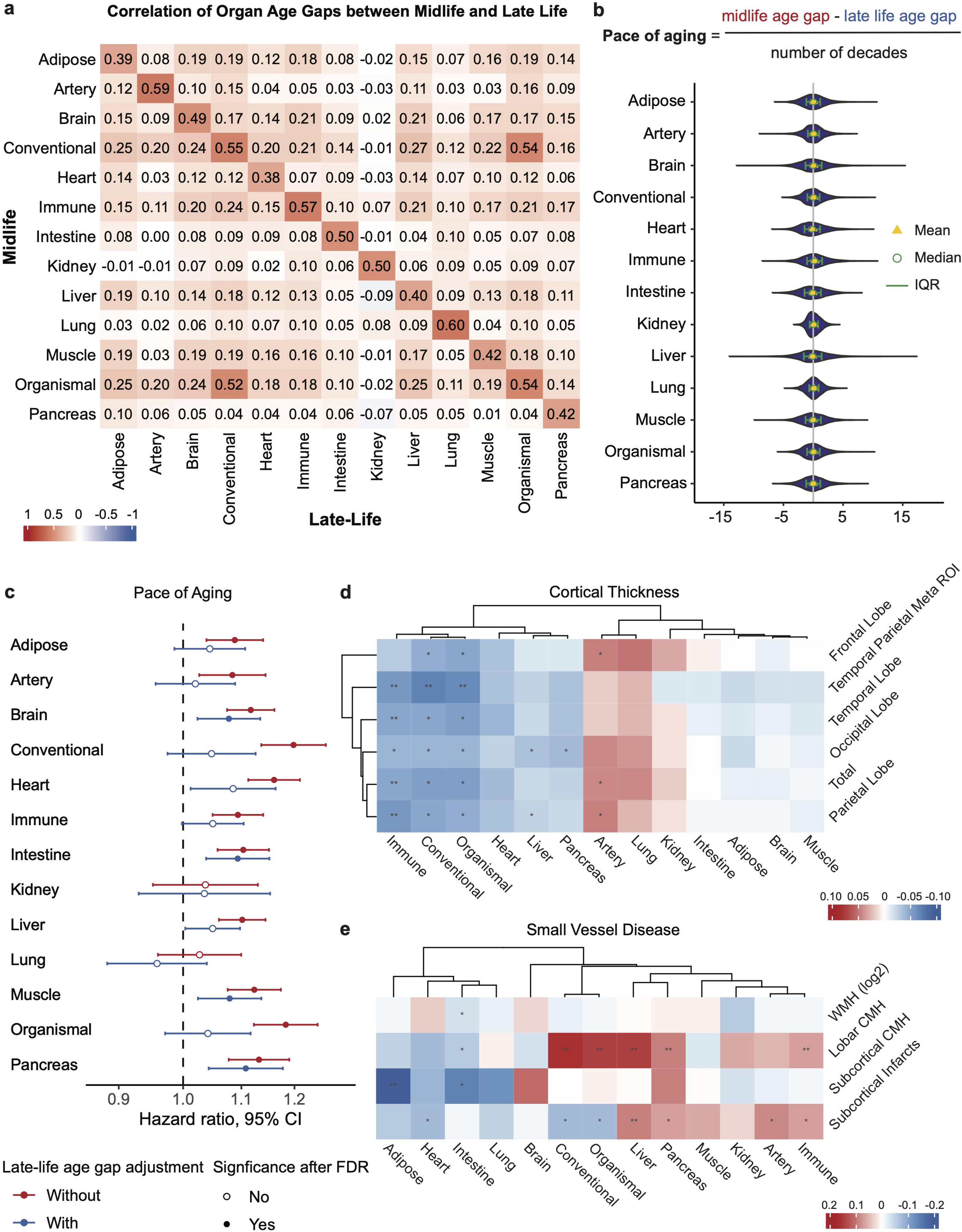
Mid- to late-life *pace of organ aging* was associated with incident dementia and MRI-defined cortical thickness and cerebral small vessel disease. All models adjusted for chronological age, sex, a composite variable of race and study center, education, body mass index (BMI), kidney function (eGFR), current smoking status, history of diabetes and hypertension, and *APOE*ε4 status. **a.** The heatmap shows the correlations of the organ age gaps within the same organ system between midlife and late life. Darker color indicates stronger correlation. **b.** The violin plot depicts the distribution of *pace of organ aging*. **c.** The forest plot shows the hazard ratio (HRs) between *pace of organ aging* and incident dementia before adjusting for late-life organ age gaps (red) and after adjusting for late-life organ age gaps (blue). **d.** The associations between *pace of organ aging* and MRI-derived cortical thickness in 5 brain regions after adjusting for late-life organ age gaps. Darker blue color indicates stronger association between faster *pace of organ aging* and lower critical thickness. **d.** The associations between *pace of organ aging* and 4 MRI-derived small vessel disease measures. Darker red color indicates stronger associations between faster *pace of organ aging* and higher white matter hyperintensity (WMH, log2-transformed) or presence of cerebral microhemorrhage (CMH)/infarcts. ** indicates statistical significance at FDR level. * indicates statistical significance at nominal level.

We first related *pace of aging* from mid- to late life to incident dementia after the late-life visit (Visit 5, N = 3,725, case = 556) over a median 6.5-year follow-up period. After adjusting for demographic and physiological variables, faster *paces of aging* in all organ systems, except for the kidney and the lung, were associated with higher risks of incident dementia (FDR p<0.05, **Figure 4C, Supplemental Table 23**). Next, we tested whether the association between *pace of aging* and dementia risk was primarily driven by the resulting larger organ-specific age gaps in late life by additionally adjusting for the corresponding late-life organ-specific age gaps. *Pace of aging* in the brain (HR=1.08; 95% CI: 1.02, 1.13), intestine (HR=1.09; 95% CI: 1.04, 1.15), muscle (HR=1.08; 95% CI: 1.02, 1.14), and pancreas (HR=1.11; 95% CI: 1.04, 1.18) remained significant (FDR p<0.05, **Figure 4C, Supplemental Table 23**), suggesting that the rate at which a person biologically ages in these organ systems is associated with dementia risk independent of the biological age they end up with. These results remain largely unchanged when we removed 98 cases and 60 non-cases with follow-up < 3 years.

Faster *pace of immune aging* was most strongly associated with lower MRI-defined cortical thickness in total brain, lobar regions, and the temporal parietal meta ROI after adjusting for the corresponding age gap in late life (FDR p < 0.05, N = 1,179, **Figure 4D**, **Supplemental Table 24**, **Supplemental Figure 5**), whereas faster *pace of liver aging* was most strongly associated with lobar cerebral microhemorrhages (case/N=211/1179) and subcortical infarcts (case/N=219/1179). Unexpectedly, faster *pace of adipose aging* was associated with lower odds of deep cerebral microhemorrhages (case/N=44/11479, **Figure 4E, Supplemental Table 25-26**).

### Proteins in circulation drive organ pace of aging and increase disease risk

Next, we identified circulating proteins and associated pathways that may contribute to dementia risk by accelerating the *pace of organ aging*. Specifically, we performed proteome wide association analysis (PWAS) to relate levels of 4,955 plasma proteins measured at midlife (Visit 2) to the *pace of aging* in 7 organ systems (brain, heart, immune, intestine, liver, muscle and pancreas) that showed associations with dementia risk and brain structure. To avoid the same protein predictor being part of the *pace of aging* simultaneously, we recalculated the *pace of aging* between Visit 3 (initiated three years after Visit 2) and Visit 5. The Pearson correlations between *pace of aging* calculated using Visit 2 age gaps and Visit 3 age gaps ranged from 0.55 (muscle) to 0.72 (heart, **Supplemental Figure 6**).

After adjusting for a composite variable of race and study center, education, body mass index (BMI), estimated glomerular filtration rate (eGFR), current smoking status, history of diabetes, hypertension, cancer, and cardiovascular disease (i.e., coronary heart disease, stroke, or heart failure), 230, 39, 134, 24, 554, 9, and 901 proteins were associated with subsequent *pace of aging* in the brain, heart, immune system, intestine, liver, muscle and pancreas, respectively (FDR p < 0.05, **Figures 5A, Supplemental Tables 27-33**). Among these proteins, TNFRSF1B, a receptor for the cytokine TNFα, and C6, a member of the multiprotein membrane attack complex (MAC), were associated with *pace of aging* in 5 of 7 organs. Another 11 proteins were significant for 4 organs (**Figure 5B, Supplemental Figure 7A**). Interestingly, some proteins showed discordant associations with pace across organs. For example, leptin (LEP), an adipose-specific protein, was associated with faster *pace of brain aging* but with slower *pace of aging in the heart, liver, and pancreas* (**Figure 5A**).

**Figure 5.**
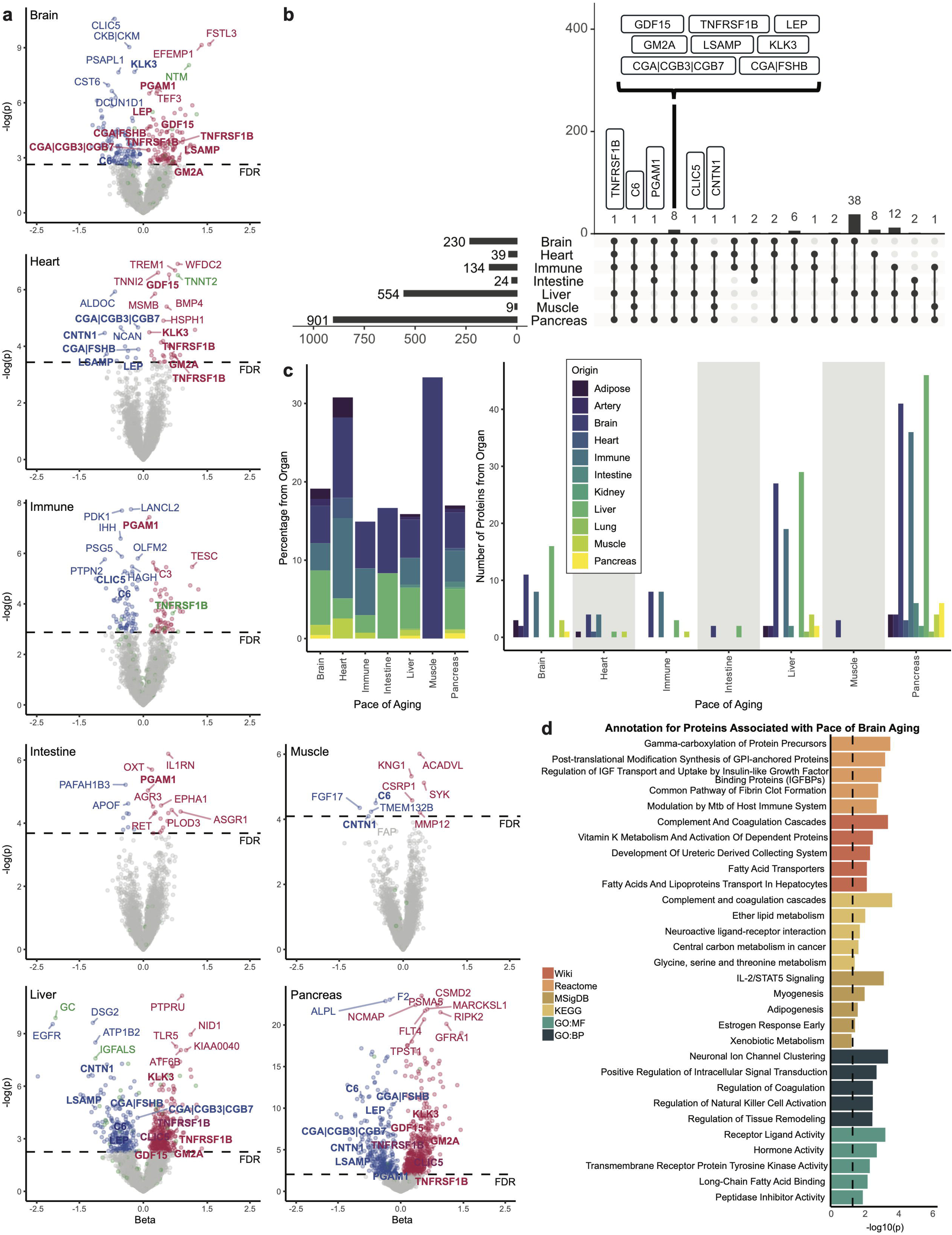
Circulating proteins were associated with *pace of organ aging.* **a.** The volcano plots shows the associations between 4,955 proteins (measured at Visit 2) and subsequent *pace of organ aging* (between Visits 3 and 5) in the brain, heart, immune system, intestine, liver, muscle and pancreas. Blue color indicates proteins that decelerated *pace of organ aging* with FDR p<0.05. Red color indicates proteins that accelerated *pace of organ aging* with FDR p<0.05. Green color indicates proteins specific to the organ of which *pace of aging* was measured. Bold font indicates proteins significantly associated with 4 or more *pace of organ aging*. The linear regression models adjusted for chronological age, sex, a composite variable of race and study center, education, body mass index (BMI), kidney function (eGFR), current smoking status, history of diabetes, hypertension, cardiovascular disease (i.e. coronary heart disease, stroke, or heart failure) and cancer, and *APOE*ε4 status. **b.** The UpSet plot shows the number of proteins significantly associated with *pace of aging* in different combinations of brain, heart, immune system, intestine, liver, muscle, and pancreas. Gene symbols for proteins associated with 4 or more *pace of aging* are annotated. **c.** The bar graphs depicts the relative (left) and absolute (right) number of proteins specific to 11 organ systems that were associated with *pace of organ aging* in the brain, heart, immune system, liver, muscle, and pancreas. **d.** The top 5 pathways or biological function for proteins significantly associated with *pace of brain aging* (FDR p<0.05) from Reactome,^41^ WikiPathways (Human),^42^ KEGG 2021 (Human),^43^ MSigDB 2020 Hallmarks,^44^ GO Biological Process 2025, and GO Molecular Function 2025.^45,46^

Proteins originating from all or nearly all organ systems were identified to be associated with *pace of brain, liver, and pancreas aging*, whereas only brain- and/or liver-specific proteins were associated with *pace of intestine and muscle aging* (**Figures 5C**). Except for the liver, few of the significant proteins were specific to the organ for which pace was being examined, suggesting a pattern of non-autonomous signaling from other organs or systemic factors may constitute a catalyst for accelerated organ aging. Results of enrichment analyses for the significant proteins associated with *pace of aging* in brain, heart, immune system, liver, and pancreas are summarized in **Figure 5D, Supplemental Figure 7B, and Supplemental Tables 34-38**. Lipid metabolism pathways were enriched among proteins associated with *pace of brain and liver aging*, whereas proteins associated with *pace of brain, liver, and pancreas aging* were enriched for complement and coagulation pathways. Proteins associated with *pace of heart and pancreas aging* were enriched for organ-specific biological pathways, e.g., muscle contraction and insulin-like growth factor related pathways, respectively. On the other hand, proteins associated with accelerated *pace of immune aging* were enriched for a broad set of biological processes, many of which are implicated in cancer.

### Proteins associated with pace of aging are causally linked to neurologic diseases

Of the 13 proteins associated with *pace of aging* in at least 4 organs, 8 proteins had *cis-*pQTLs identified in ARIC,^22^ we used in Mendelian randomization (MR) to examine their potential causal role in accelerating or decelerating *pace of organ aging*. The outcome summary statistics were derived from a GWAS of the *pace of brain, heart, immune, intestine, liver, muscle, and pancreas aging* between Visit 3 and Visit 5 (**Supplemental Data**). Consistent with the associative analysis, MR analyses demonstrated that increased TNFRSF1B accelerated the *pace of aging in the brain, immune system, muscle, and pancreas* among White participants (nominal p<0.05). On the other hand, MR analysis of C6 showed a putative protective causal effect on the *pace of pancreas aging* in Black participants (**Figure 6, Supplemental Table 39**). Although TNFRSF1B is an immune-specific protein, it contributed minimally to the prediction of immune age.^6^ The only other organ-specific protein, CNTN1 (brain-specific), did not show a putative causal relationship with *pace of aging in the brain*. All other 6 proteins were not part of the organ-age estimation. Taken together, these potential causal relationships were not solely driven by the proteins’ organ specificity.

**Figure 6.**
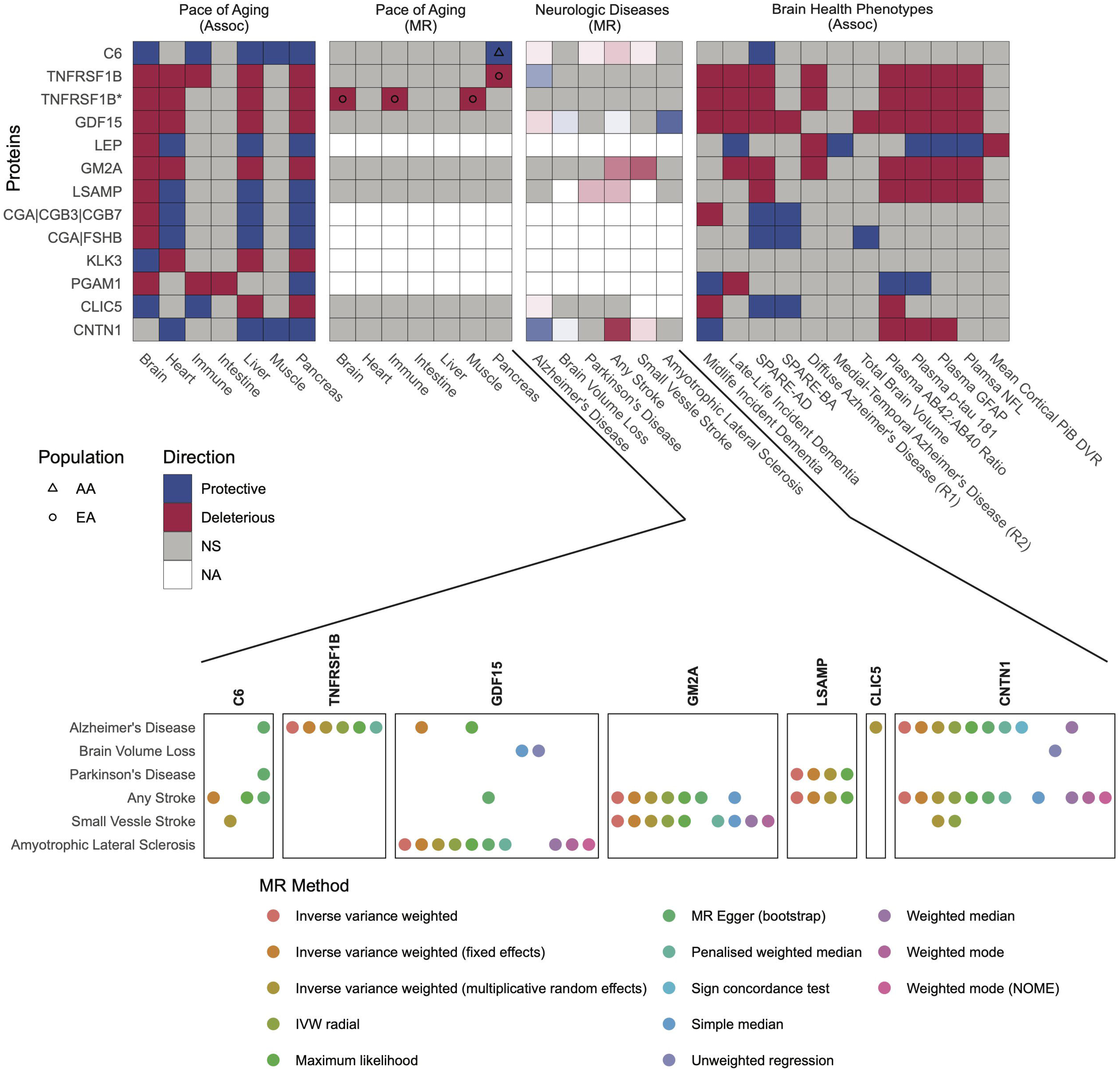
Proteins associated with *pace of organ aging* in 4 or more organ systems were causally linked to *pace of organ aging* and neurologic disease. The 1^st^ panel on the top shows the direction of associations between the 13 proteins significantly associated with 4 or more *pace of aging* in the brain, heart, immune system, intestine, liver, muscle, and pancreas and the *pace of aging* in those organs. Of the 13 proteins, 8 proteins had *cis-*pQTLs identified in ARIC.^21^ Mendelian randomization (MR) analyses linking the 8 proteins and *pace of organ aging* are shown in the 2^nd^ panel on the top. These proteins were also causally linked to various neurologic diseases using two-sample MR (3^rd^ top panel). Darker color in this panel indicates a greater number of two-sample MR methods suggesting a causal link between the proteins and neurologic disease (bottom panel), The 4^th^ top penal shows the direction of associations between the 13 proteins and various brain health phenotypes in ARIC and BLSA.

In two-sample MR analyses that related each of the 8 *pace of organ* aging proteins to neurologic diseases using published GWAS summary statistics,^23–27^ we found several putative causal associations that were directionally opposite of our associative analyses (**Figure 6, Supplemental Table 40**). Although midlife TNFRSF1B was associated with a faster pace of brain aging and C6 with a slower *pace of brain aging*, MR results suggested they were protective against and deleterious to AD, respectively. Interestingly, midlife GDF15’s association with a faster *pace of brain aging* aligns with the putative pathogenic causal effect of this protein in AD (confirmed by previous studies^28,29^); however, MR also suggested plasma GDF15 is protective against brain volume loss, stroke, and Amyotrophic Lateral Sclerosis (ALS). More consistent findings were detected for limbic system-associated membrane protein (LSAMP), a protein involved in axon guidance and neuronal growth in the developing limbic system, and GM2 ganglioside activator (GM2A), a protein responsible for breaking down neurotoxic GM2 ganglioside. Plasma levels of both proteins were associated with a faster *pace of brain aging*, and MR analyses supported the deleterious effects on brain volume loss and/or stroke.

Analyses relating the 13 *pace of organ aging* proteins to dementia risk and MRI-, PET-, and plasma-defined brain health phenotypes in ARIC and the Baltimore Longitudinal Study of Aging (BLSA), a study of normative human ageing established in 1958 with continuous enrollment to date,^30^ (**Figure 6, Supplemental Tables 41-42**) largely aligned with each protein’s *pace of brain aging* association. TNFRSF1B showed a consistent and robust association with higher dementia risk in mid- and late life, as well as pathogenic ADRD biomarker levels. The association between C6 and brain health was minimal. LSAMP and GM2A showed deleterious associations with SPARE-AD, an MRI-based AD-like pattern of brain atrophy, and ADRD biomarkers, consistent with their associations with faster *pace of brain aging* and their putative causal role in neurologic disease (MR results).

## Discussion

Our study extends previous studies linking organ aging and dementia by demonstrating that organ systems other than the brain, including the heart, immune system, intestine, liver, muscle, and pancreas, may also be important determinants of cognitive decline and future dementia risk, individually and/or synergistically. Leveraging longitudinal proteomic measurement, we demonstrate that a faster *pace of brain, intestine, muscle, and pancreas* aging over a multi-decade period from midlife to late life is associated with greater subsequent risk of dementia, independent of the organ age in late life. We demonstrate that this is not only the case for dementia, but also for the MRI-defined measures of neurodegeneration and cerebral small vessel disease, which underlie cognitive decline and dementia. Taking a proteome-wide approach to understand the systemic factors that accelerate or decelerate the *pace of organ aging*, we found that organ non-autonomous proteins were most strongly associated with *pace of organ-specific aging* during the decades spanning mid- to late-life. This was especially true for the rates of brain, heart, liver, and pancreas aging, which all have broad proteomic signatures largely independent – in terms of the origin of the protein – from the aging organ. We identified a set of proteins associated with the *pace of aging* of four or more dementia-related organ systems and used two-sample Mendelian randomization to find causal support for the link between plasma TNFRSF1B and accelerated *pace of brain, immune system, muscle, and pancreas aging*.

To our knowledge, our study is the first study to demonstrate that the longitudinal *pace of organ aging* is associated with dementia risk and brain health independent of late-life organ age. We are also among the first to show that advanced age in multiple organs is associated with dementia risk. Findings by Oh and colleagues^12^ that mortality increased supra-additively with increasing number of extremely aged organs corroborated the supra-additive increase in dementia risk we observed. Together, these findings are consistent with the theory that multiple perturbations to a complex system do not act additively but may instead exhibit synergistic effects on age-related conditions, such as dementia.^31^ In particular, the combinations of advanced brain and heart age and advanced brain and muscle age in midlife may make individuals especially vulnerable to dementia risk. These findings support the robust link between heart health, physical frailty, and dementia risk, and highlight the role that interventions targeting these systems may have in delaying dementia onset.^32^

We found support for the crosstalk among organ systems in the human body, as mediated by the circulating proteome. For example, our results indicate that TNFRSF1B, an immune system-specific protein, may causally increase the *pace of aging* in the brain, muscle, and pancreas. Despite being mechanistically associated with a faster pace of aging across multiple organs, MR analyses did not support TNFRSF1B’s pathogenic role in neurologic disease. On the other hand, complement factor C6, which was associated with a reduced *pace of pancreatic aging*, appeared to increase risk for stroke in MR analyses. Interestingly, although CNTN1 was associated with a slower *pace of aging in heart, liver, muscle, and pancreas*, and MR supported its protective role in AD, our findings suggest CNTN1 may also increase stroke risk. Because multiple MR methods were applied, the discrepancy could be due to differing sensitivity to violations of assumptions by each MR method. Competing risk (the diagnosis of one disease precludes the diagnosis of the other in MR)^33^ and diverging effects in midlife (time frame for *pace of organ aging*) versus late life (time frame for MR of neurologic diseases) may be other explanations of the discrepant results. More consistently, GM2A and LSAMP, both associated with a faster *pace of brain aging* and many brain health phenotypes, were also mechanistically linked to increased risk of stroke and Parkinson’s disease. These findings are supported by previous results linking GM2A and LSAMP to reduced neurite integrity^34^ and dementia risk,^35^ respectively.

Our study possesses many strengths, including multi-decade and biracial longitudinal proteomic measurement, but it is not without limitations. First, the determination of dementia subtypes based on available measures in ARIC is unreliable. Therefore, we did not investigate different dementia etiologies. In one study on brain organ age only, the association of extremely older brain age gap with vascular dementia was smaller than with AD.^13^ Future studies are needed to further elucidate whether different organs play more important roles in different dementia subtypes. Second, the sample size for the GWAS of *pace of organ aging* was small. Therefore, we may not have sufficient power to detect the causal effects of some of the proteins on *pace of organ aging* in our MR analyses. Moreover, it is generally not recommended that instrument discovery (*cis-*pQTL analysis) and MR are performed in the same population due to the winner’s curse.^36^ However, due to limited published *cis*-pQTL results in the midlife population and studies with longitudinal proteomics to perform GWAS of *pace of organ aging*, our options were limited. Future studies are encouraged to replicate and extend our results in larger, more diverse cohorts. Despite these limitations, our results highlight the potential of proteomic aging measures in multiple organs across time as dynamic indicators and potential drivers of dementia risk. Ultimately, understanding how different organs age together—or diverge—may open new opportunities for early prevention and precision interventions to promote brain health and longevity.

## Methods

### Study cohort and design

The Atherosclerosis Risk in Communities (ARIC) study is an ongoing community-based cohort that enrolled participants 45-64 years from 4 communities across the United States in 1987-1989 (Visit 1): Washington County, MD; Forsyth County, NC; northwestern suburbs of Minneapolis, MN; and Jackson, MS.^15^ Proteomics measured using the SomaScan assay^®^ (Version 4.0, SomaLogic, Inc) were available for Visit 2 (1990-1992) and Visit 5 (2011-2013), which were considered the baseline for our midlife and late-life analyses, respectively. For midlife, of the 14,348 participants who returned to Visit 2, we excluded 91 (<1%) participants who were of non-Black and non-White races and Black participants in the Minnesota and Maryland sites due to small sizes. We also excluded 2,464 (17%) participants who did not have proteomics measures. We further excluded 191 (1%) participants who had missing covariate values and 7 (<1%) participants who had a dementia diagnosis (described below) before Visit 2 or missing diagnosis at follow-up, resulting in a sample of 11,595 participants. Similarly, for late life, of the 6,538 participants who returned to Visit 5, we excluded 43 (<1%) participants from the minor race-center groups, 1,015 (16%) who had no proteomics measure, 496 (8%) who had missing covariates, 405 (6%) who had a dementia diagnosis or missing diagnosis before Visit 5, and 292 (4%) who had missing diagnosis at follow-up, resulting a sample of 4,287 participants.

### Dementia adjudication

The detailed adjudication has been described elsewhere.^37,38^ For participants who attended Visits 5, 6, and 7, dementia was diagnosed based on an algorithm based on the National Institute on Aging–Alzheimer’s Association (NIA-AA) workgroups^39,40^ and D*iagnostic and Statistical Manual of Mental Disorders, 5th Edition (DSM-5)*. The algorithm utilized the Visit 5 in-person evaluations of a 10-test neurocognitive battery, the Mini-Mental State Exam (MMSE), the Clinical Dementia Rating (CDR) scale, the Functional Activities Questionnaire (FAQ), and the change scores of delayed word recall (DWR), digit symbol substitution (DSS), and word fluency test (WFT) since Visit 2. The 10-test battery included DWR, DSS, WFT, incidental learning (ILR), animal naming score (ANS), logical memory test (LMT), trail making test A (TMTA) and B (TMTB), digit span backwards (DSB), and Boston naming test (BNT). An expert committee confirmed or overruled the algorithmic diagnoses. The date of diagnosis was set to the date of visits unless a hospital discharge record indicated an earlier date.

For participants who did not attend Visits 5-7 including due to death, dementia diagnoses prior to June 2015 were ascertained by the Telephone Interview for Cognitive Status–Modified (TICSm) score, the informant-completed CDR and FAQ (in the absence of TICSm). After June 2015, six-item screener (SIS) and eight-item interview to differentiate aging and dementia (AD8) were used to ascertain the diagnosis in the absence of TICSm, CDR, and FAQ. The date of the diagnosis was set to the earliest date when any of the above instruments detected dementia. If a participant had none of the above assessments, the hospital discharge records, and death certificates were used to determine the diagnosis. The date of diagnosis was set to 180 days before the hospital discharge date or date of death.

### MRI-based brain structure

Cortical thickness, white matter hyperintensity, microhemorrhages, and infarcts were all measured using 3T MRI scanners (Maryland: Siemens Verio; North Carolina: Siemens Skyra; Minnesota: Siemens Trio; Mississippi: Siemens Skyra). At Visit 5, participants who were diagnosed with mild cognitive impairment (MCI) or had a prior MRI measurement and an age-and study site-stratified random sample of cognitively unimpaired were invited to complete the MRI scans. T1-Weighted Magnetization Prepared Rapid Acquisition Gradient-Echo (MPRAGE), T2 Fluid-Attenuated Inversion Recovery (FLAIR), and gradient recalled echo T2-weighted imaging (T2*GRE) were sequences used for the measurement of cortical thickness, white matter hyperintensity and infarcts, and microhemorrhages, respectively.^41^

The cortical thickness was assess using the Freesurfer atlas.^42^ We pre-specified the regions of interest (ROIs) to include: (1) the frontal lobe (caudal anterior cingulate, caudal middle frontal, lateral orbitofrontal, medial orbitofrontal, paracentral, pars opercularis, pars orbitalis, pars triangularis, precentral, rostral anterior cingulate, rostral middle frontal, superior frontal, and frontal pole regions); (2) temporal lobe (bankssts, entorhinal, fusiform, inferior temporal, middle temporal, parahippocampal, superior temporal, temporal pole, and transverse temporal regions); (3) occipital lobe (cuneus, lateral occipital, lingual, and pericalcarine regions); (4) parietal lobe (inferior parietal, isthmus cingulate, postcentral, posterior cingulate, precuneus, superior parietal, and supramarginal regions); (5) temporal parietal meta ROI (entorhinal, fusiform, inferior temporal, middle temporal, and precuneus regions);^43^ (6) total brain.

White matter hyperintensity was measured using an algorithm developed at the Mayo Clinic in Rochester, Minnesota.^44^ Infarcts and microhemorrhages were identified, counted, and measured by a trained imaging technician and confirmed by a radiologist. Subcortical infarcts were defined as T2 FLAIR lesions with central hypointensity >3 mm and hyperintensity ≤20 mm in maximum dimension located in the caudate, lenticular nucleus, internal capsule, thalamus, brainstem, deep cerebellar white matter, centrum semiovale, or corona radiata. Lobar cerebral microhemorrhages were counted in the periventricular and subcortical regions of the 4 lobes. Deep cerebral microhemorrhages were counted in the internal and external capsule, deep grey and white, corpus callosum, anterior corpus callosum, posterior corpus callosum, and central regions.

### Organ-specific age gap prediction

We used the algorithm developed by Oh and colleagues that map plasma proteins to organ systems based on human organ bulk RNA sequencing (RNA-seq) data from Genotype-Tissue Expression (GTEx) project.^6^ Genes expressed at least 4 times higher in one organ than any other organs are considered specific to that organ. Separate machine learning models of chronological age were trained for the 11 organ systems we examined in this paper, using the organ-specific proteins and sex. We applied the original weights from Oh and colleagues’ algorithm to the ARIC data to obtain predicted plasma proteomic based age, using the “organage” Python package. The age gaps were then obtained based on the distribution of predicted age in ARIC. This is achieved by fitting visit-specific lowess curves of predicted age for each organ system versus chronological age and calculating the difference between the predicted age and the lowess curve. This procedure was implemented separately for Visits 2 (midlife), 3, and 5 (late life). We calculated the pace of aging by subtracting the age gaps at Visit 2 (for the analysis with dementia risk and MRI-based brain structures) or 3 (for the proteome wide association analysis) from the age gaps at Visit 5. Pace of aging was standardized by the time lapsed (in decades) between the two visits (Visit 2/3 and 5).

### Covariates

Participants self-reported sex (male/female), race (Black/White) and education level (less than high school/high school, general education diploma or vocational school/some college education) at Visit 1. A composite variable of race and study center was used in statistical analyses because Jackson, MS center recruited Black participants only. All other covariates were measured at Visit 2 and Visit 5 for midlife and late-life analyses, respectively. BMI was calculated using height and weight. Kidney function was measured by estimated glomerular filtration rate (eGFR) calculated using serum creatinine, age, and sex in the CKD-EPI 2012 Equation.^45^ Current smoking status was self-reported at each visit. Hypertension was defined as present if a systolic blood pressure >140 mmHg, a diastolic blood pressure >90 mmHg, or use of hypertensive medication. Diabetes at Visit 2 was defined as present if a fasting glucose ≥126 mg/dL, non-fasting glucose ≥200 mg/dL, current use of diabetes medication, or self-reported physician diagnosis. Diabetes at Visit 5 was defined as present if hemoglobin A1C value>=6.5% or using medication for diabetes or self-reported physician diagnosis. *APOE* was genotyped using the TaqMan assay and coded as 0, ≥1, or missing ε4 alleles.

### Statistical analyses in ARIC

We used Cox proportional hazard models to examine the associations of (a) age gaps in midlife and late life, (b) pace of aging, (c) number of organs with abnormal age gaps, and (d) the combination of abnormal brain age gap and abnormal age gap in another organ system with dementia. The proportional hazard assumption and linearity assumption were checked and confirmed by the Schoenfeld residuals and Martingale residuals respectively (data not shown). We used linear regression models and logistic regression models to examine the associations of (a)–(d) with MRI-defined cortical thickness and cerebral small vessel disease, adjusting for the same covariates. All models were adjusted for chronological age, sex, a composite variable of race and study center, education, BMI, kidney function, current smoking status, history of diabetes and hypertension, and *APOE*ε4 status.

Linear regression models were used for the proteome wide association analysis between plasma proteins at Visit 2 and the pace of aging between Visit 3 and Visit 5. In addition to the above-mentioned covariates, we further included history of cardiovascular disease (i.e. coronary heart disease, stroke, or heart failure) and cancer, both are associated with plasma proteins and could influence pace of organ aging. All the above analyses were performed in R (version 4.5.0).

### Pathway and biological function enrichment

We performed enrichment analysis for proteins significantly associated with pace of aging in brain, heart, immune, liver, and pancreas (FDR p <0.05) using Enrichr (https://maayanlab.cloud/Enrichr/).^46^ We excluded intestine and muscle because they had <30 significant proteins. We used all 4,955 proteins measured in ARIC as the background. We extracted the top 5 pathways or biological functions from Reactome,^47^ WikiPathways (Human),^48^ KEGG 2021 (Human),^49^ MSigDB 2020 Hallmarks,^50^ GO Biological Process 2025, and GO Molecular Function 2025.^51,52^

### GWAS for organ pace of aging and Mendelian randomization

Genotyping ARIC has been described elsewhere and was imputed to the TOPMed reference panel (Freeze 5b).^53,54^ The GWAS was performed separately in White and Black participants using PLINK 2.0. We excluded SNPs with imputation quality R^2^ < 0.8, call rates < 95%, Hardy-Weinberg equilibrium p-values < 10^-8^, or minor allele frequencies < 1% and participants with > 5% missing genotypes, resulting in 7,793,990 SNPs for 3,128 White participants and 13,814,458 SNPs for 610 Black participants who had pace of aging measured between Visit 3 and Visit 5. We used linear additive regression models adjusting for age, sex, study center, and 10 principal components of ancestry.

Mendelian randomization (MR) analyses to explore potential causal proteins for accelerated or decelerated pace of organ aging were performed using the R-based FUSION pipeline^55^ that leverages previously published ARIC *cis-*pQTLs^22^ prediction models along with the above GWAS results. This method allows the use of large numbers of potentially correlated *cis-*pQTLs as instrumental variables.^22^ Analyses were performed using R version 4.4.0.

For MR analyses to explore potential causal proteins for neurologic diseases, we used the same *cis-*pQTLs found in ARIC as instruments but performed clumping using ± 500kb window and r^2^ <0.05 using PLINK 2.0. GWAS summary statistics for the neurological diseases were obtained from published data in independent cohorts.^23–27^ Multiple two-sample MR methods (namely, inverse variance weighted [IVW] methods, maximum likelihood, MR Egger, penalised weighted median, sign concordance test, simple median, weighted median, weighted mode, and unweighted regression) were applied using the “TwoSampleMR” package in R^56^.

### Association analyses for brain health phenotypes

Associations between the 13 proteins associated with ≥4 pace of organ aging and midlife and late-life incident dementia were examined in ARIC using the same method described previously.^57^ Incident dementia cases in midlife and late life were identified during the same time period and using the same definition as the main analyses in the present paper. Same covariates were adjusted in the Cox proportional hazard models.

Associations between the proteins and other brain health phenotypes were examined in the BLSA.^30^ All phenotypes were measured concurrently with the proteomics measured using SomaScan assay^®^ (Version 4.1, SomaLogic, Inc).^58,59^ We examined total brain and several brain atrophy patterns derived from MRI. SPARE-AD (Spatial Patterns of Abnormality for Recognition of Early Alzheimer’s Disease) captures atrophy patterns that are consistent with Alzheimer’s disease.^60^ SPARE-BA (Spatial Pattern of Atrophy for Recognition of Brain Aging) reflects general aging.^61^ R1 and R2 are two latent dimensions that represents diffuse AD brain atrophy and medial temporal lobe AD atrophy, respectively.^62^ We also examined several plasma-based AD-specific and non-specific biomarkers. Lower plasma Aβ42/40 ratio predicts PET Aβ-positive status and changes prior to symptoms of AD. Higher plasma p-tau 181 indicates AD-specific soluble tau pathology. Neurofilament light chain (NFL) and glial fibrillary acidic protein (GFAP) are non-AD specific markers of neuronal injury and reactive astrocytosis, respectively.^16^ Lastly, we examined mean cortical Aβ burden using the average distribution volume ratio (DVR) in cingulate, frontal, parietal (including precuneus), lateral temporal, and lateral occipital cortical regions, excluding the sensorimotor strip from PiB PET imaging.^63^ All phenotypes were analyzed continuously using linear regression models adjusted for age, sex, race, education, eGFR, comorbidity index of obesity, hypertension, diabetes, cancer, ischemic heart disease, chronic heart failure, chronic kidney disease, and chronic obstructive pulmonary disease, and *APOE*ε4 status.

## Acknowledgement

We thank Dr. Basilio Cieza Huaman, Dr. Jarod Rutledge, and Mr. Alex Boches for their contribution to the study design and feedback at the various stages of the project.

## Competing interests

J.Coresh was a scientific advisor to Soma Logic until November, 2023. K.A.W. is an Associate Editor for Alzheimer’s & Dementia: The Journal of the Alzheimer’s Association, Alzheimer’s & Dementia: Translational Research and Clinical Interventions (TRCI), and on the Editorial Board of Annals of Clinical and Translational Neurology. K.A.W. is on the Board of Directors of the National Academy of Neuropsychology. K.A.W. has given unpaid presentations and seminars on behalf of SomaLogic. T.W.C. is a co-founder and scientific advisor of Vero Biosciences and Teal Rise.

## Funding statement

The Atherosclerosis Risk in Communities Study is carried out as a collaborative study supported by National Heart, Lung, and Blood Institute contracts (75N92022D00001, 75N92022D00002, 75N92022D00003, 75N92022D00004, 75N92022D00005). The ARIC Neurocognitive Study is supported by U01HL096812, U01HL096814, U01HL096899, U01HL096902, and U01HL096917 from the NIH (NHLBI, NINDS, NIA, and NIDCD). The authors thank the staff and participants of the ARIC study for their important contributions. SomaLogic Inc. conducted the SomaScan assay^®^ in exchange for the use of ARIC data. This work was supported in part by NIA Intramural Research Program (F.L., C.J., M.R.D., and K.A.W.) and NIH/NHLBI grant R01 HL134320. A.P. is supported by NIH/NIA grant R21 AG079242. P.S. and Z.R-H. are supported by the German Research Foundation (DFG) Project-ID 530592017 SCHL 2292/3–1. T.W.C is supported by the Phil and Penny Knight Initiative for Brain Resilience. S.W. is supported by the National Center for Advancing Translational Sciences grant UM1TR004405. The contributions of the NIH author(s) are considered Works of the United States Government. The findings and conclusions presented in this paper are those of the author(s) and do not necessarily reflect the views of the NIH or the U.S. Department of Health and Human Services.

## Data availability statement

Pre-existing data access policies for each of the parent cohort studies specify that research data requests can be submitted to each steering committee; these will be promptly reviewed for confidentiality or intellectual property restrictions and will not unreasonably be refused. Please refer to the data sharing policies of these studies. Individual level patient or protein data may further be restricted by consent, confidentiality or privacy laws/considerations. These policies apply to both clinical and proteomic data.

## Notes

### Author Declarations

Ethics committee/IRB of Johns Hopkins University gave ethical approval for this work

