## Supplemental Figure for "Accelerated multi-organ proteomic aging is detectable decades before dementia onset"

**Supplemental Figures**

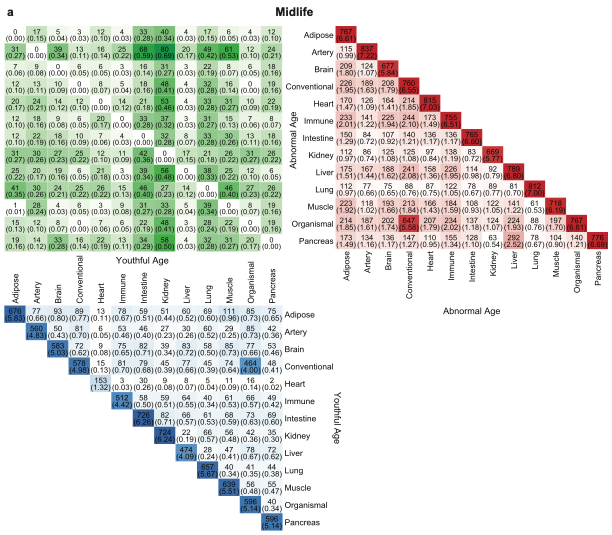

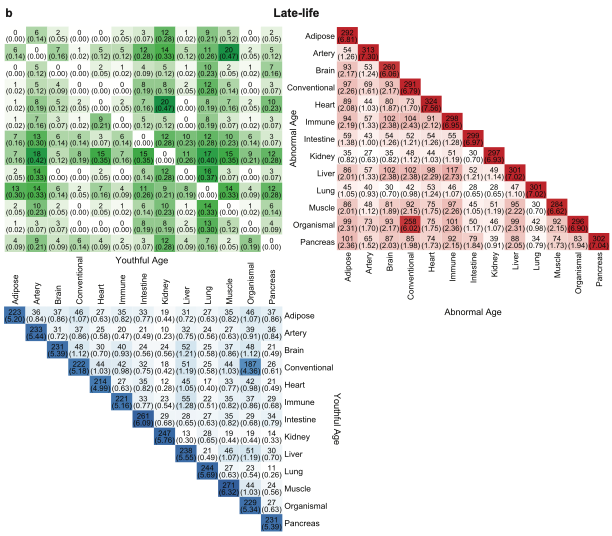

**Supplemental Figure 1.** Number of individuals with abnormal age (>1.5 SD) in one organ and youthful age (<-1.5 SD) in another organ (top left), abnormal age in two organs (top right), and youthful age in two organs (bottom left). **a.** midlife. **b.** late-life.

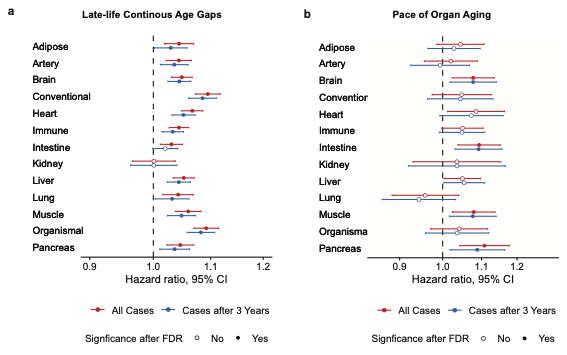

**Supplemental Figure 2.** Change in the associations with dementia risk excluding cases that occurred within 3 years from baseline, compared to dementia risk including all cases after baseline. **a.** late-life age gaps. **b.** *pace of organ aging* between midlife and late-life.

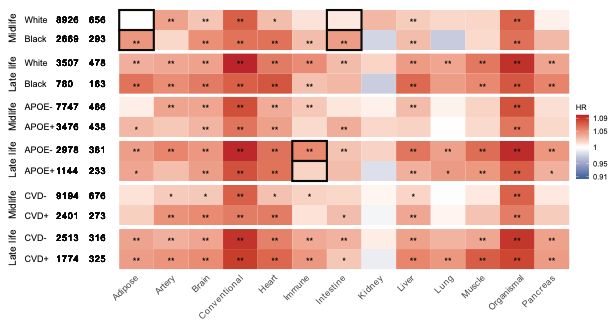

**Supplemental Figure 3.** The HRs between organ age gaps and incident dementia comparing Black and White participants in midlife (first two rows), *APOE*ε4 carriers to non-carriers (middle two rows), and participants with to without history of CVD at baseline (bottom two rows). Black boxes indicate organ age gaps that showed significantly different associations with incident dementia between groups. ** indicates statistical significance at FDR level. * indicates statistical significance at nominal level.

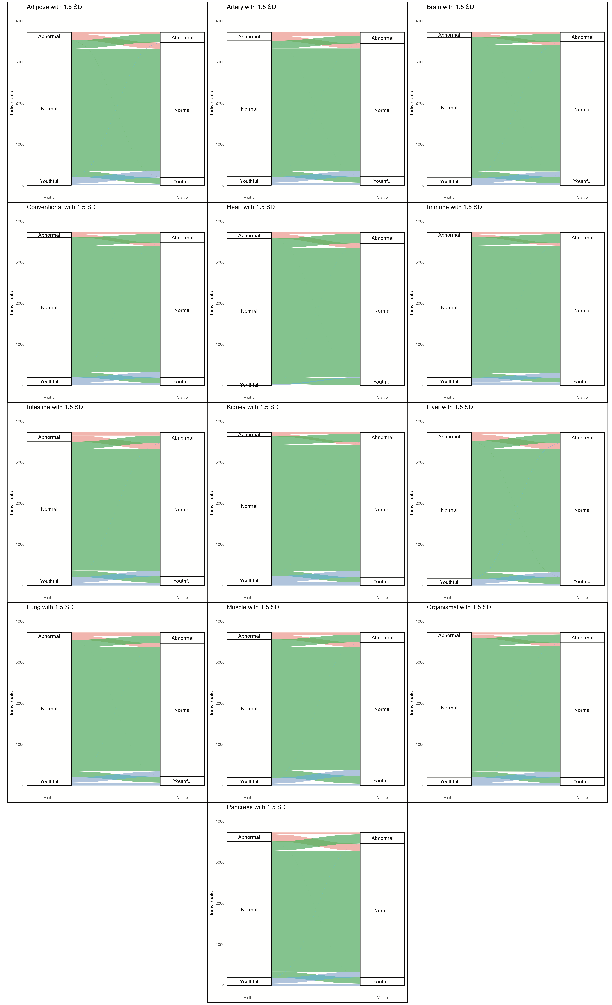

**Supplemental Figure 4.** Transitions between organ-specific age categories (abnormal, normal, and youthful) between midlife and late life.

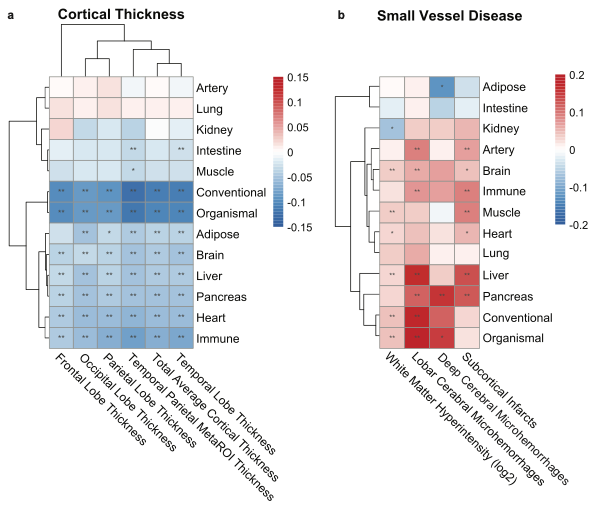

**Supplemental Figure 5. a.** The associations between *pace of organ aging* and MRI-derived cortical thickness in 5 brain regions. Darker blue color indicates stronger association between more advanced organ age and lower critical thickness. **b.** The associations between *pace of organ aging* and 4 MRI-derived small vessel disease measures. Darker red color indicates stronger associations between more advanced organ age and higher white matter hyperintensity (WMH, log2-transformed) or presence of cerebral microhemorrhage (CMH)/infarcts. ** indicates statistical significance at FDR level. * indicates statistical significance at nominal level.

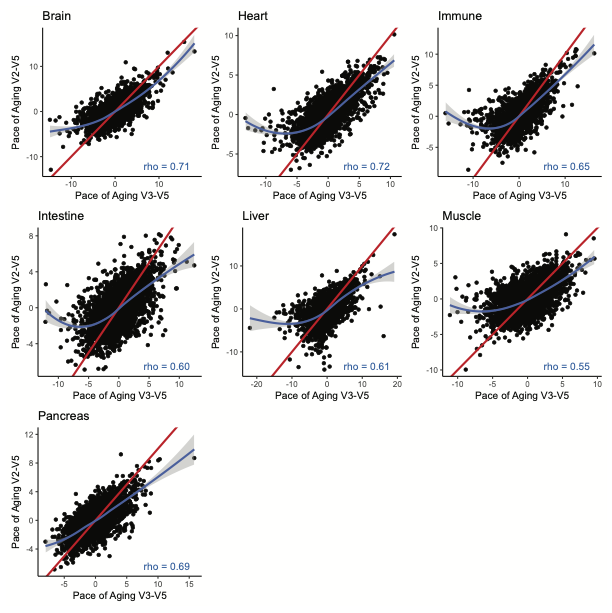

**Supplemental Figure 6.** Correlations between *pace of organ aging* between Visit 2 and Visit 5 and *pace of organ aging* between Visit 3 and Visit 5.

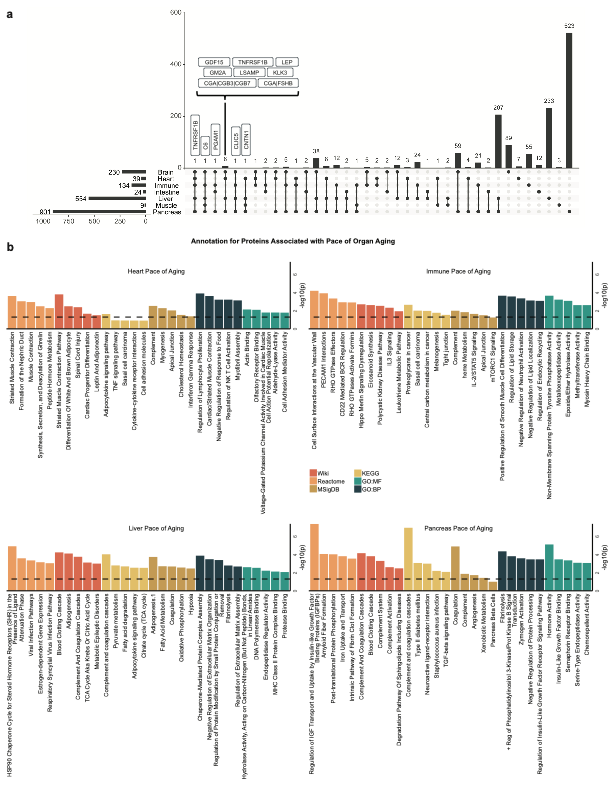

**Supplemental Figure 7. a.** The UpSet plot shows the number of proteins significantly associated with *pace of aging* in different combinations of brain, heart, immune system, intestine, liver, muscle, and pancreas. Gene symbols for proteins associated with 4 or more *pace of aging* are annotated. **b.** The top 5 pathways or biological function for proteins significantly associated with *pace of aging* in the heart, immune system, liver, and pancreas (FDR p<0.05) from Reactome,^41^ WikiPathways (Human),^42^ KEGG 2021 (Human),^43^ MSigDB 2020 Hallmarks,^44^ GO Biological Process 2025, and GO Molecular Function 2025.^45,46^
